# Exploring Gender Bias in Cardiovascular Medical Education Through Clinical Simulation

**DOI:** 10.1101/2024.05.22.24307766

**Authors:** Idris F. Ali Amghaiab, Archana Venkatesan, Matthew C. Tews, A.J. Kleinheksel

## Abstract

While it has been proven that women suffer disproportionately from cardiovascular disease-related deaths, the origins of this differential gender-based outcome remain unidentified. One possible cause is gender bias and associated discrepancies in how physicians assess the need for interventions in male versus female patients. This study aimed to identify early gender biases in cardiovascular care by assessing medical students’ management of a ruptured abdominal aortic aneurysm case presentation in male and female simulated patients. Clerkship students (n = 187) were randomly assigned to either a male or female patient with identical case presentations, simulated using high-fidelity mannequins. Minutes passed until point-of-care ultrasound (POCUS) usage served as a surrogate for diagnostic reasoning, while minutes passed until surgery consultation call served as a surrogate for successful intervention.

Two-way ANCOVA of time to surgery call and POCUS use showed no significant interaction between student and patient gender (p=0.819). Likewise, neither patient gender (p=0.210) nor student gender (p=0.653) had an impact on ultimate correct diagnosis. However, there appeared to be an association (p=0.010) between patient and student gender in the factorial ANOVA of POCUS use, F(1,183) = 6.862 effect size 0.36. While slight in-group bias was identified within the context of imaging, students predominantly called for the correct intervention regardless of their own or their patient’s gender. It is thus unlikely that medical students develop gender biases serious enough to impact clinical outcomes during clerkship-instead, these heuristics may be formed later in training.

## Introduction

Cardiovascular disease is a leading cause of illness and mortality nationwide^1^. Yet, while it remains a leading cause of death for both men and women, women suffer from a disproportionately higher mortality rate from cardiac-related illness^2^. While sex-based differences in cardiac disease presentation may contribute to this problem, the effects of gender bias in the course of treatment may exacerbate this disparity^2-4^. The origins of this bias, however, remain unidentified.

In the presentation of cardiovascular heart disease, both men and women will present most commonly with the “typical” cardiac-related symptoms. However, women present much more frequently than men with what are often referred to as “atypical” and “non-specific” cardiac symptoms, which include but are not limited to fatigue, anxiety, and nausea^3-5^. Due to these atypical presentations of disease, women may receive different levels of emergency room care, including longer wait times and reduced rates of prophylactic treatment at discharge^3^,^6^,^7^. The erroneous belief that women are somehow protected from cardiovascular disease, or that their atypical symptoms mean they are less likely to be experiencing a cardiac event, is an important area of research. Previous studies have shown gender disparities exist in diagnostic strategies. For example, women with obstructive congenital heart disease were less likely to undergo diagnostic studies such as stress tests and diagnostic angiograms than males^8^. This bias translates to procedural management practices as well. Even after adjustment for patient medical history and presentation, women were significantly less likely than men to receive invasive coronary revascularization during follow-up after coronary artery disease^9-11^. Likewise, women underwent coronary artery bypass grafts (CABG) less frequently than males with similar baseline characteristics^12^. It has further been revealed that, in general, women undergoing cardiovascular procedures were about three years older and had slightly higher scores on illness severity indices than men^13^. These findings indicate discrepancies in how physicians assess the need for diagnostic and procedural interventions in male versus female patients with similar disease states. Naturally, these disparities in treatment translate to differential patient outcomes. For example, after controlling for confounding factors such as age, race, severity of illness, and hospital location, women were 13% more likely to die in the hospital after a heart procedure (including pacemaker implantations, stent insertions, and heart valve replacementS) than male counterparts^13^.

Standardized care practices merit the development of objective guidelines to help improve the health outcomes of women, as they have long been neglected in clinical studies and trials. Guidelines to manage heart disease have largely been based on the male population^5-7^,^14^, and implicit biases and assumptions have further contributed to worse clinical outcomes for the female population^2^,^14^. Due to these poorer outcomes, and the higher mortality rates from cardiovascular disease observed in women, the origins of gender bias in healthcare must be explored so that it can be addressed and its effects can be mitigated.

While it is difficult to pinpoint where these biases originate, medical practitioners are typically first introduced to the foundational aspects and nuances of medicine in medical school. One such modality through which students are taught clinical concepts is through the use of high-fidelity simulation activities, a practice that is quickly gaining traction among medical schools across the country. In particular, studies have demonstrated that simulated practice can improve performance outcomes in treatment of cardiac cases^15^. Despite their widespread use, it can be difficult to evaluate if case-based curriculum leads to bias that may impact learners as they progress in their medical careers. Identifying the stage in medical education during which physicians develop biases will better equip us to develop strategies to mitigate them. As such, the goal of this study is to potentially identify early biases in cardiovascular care by assessing third-year medical students’ management of a standardized ruptured abdominal aortic aneurysm (AAA) case presentation in male versus female simulated patients.

## Methods

Over two days in December 2019, 187 third-year medical students at a large public medical school in the Southeastern United States participated in a formative individual simulation case with immediate post-event debriefing during an intersession between the two six-month clerkship rotation blocks. During the simulation of a ruptured abdominal aortic aneurysm (AAA), students were randomly assigned to a high-fidelity patient mannequin of either the male or female gender, with identical case presentation, vitals, and patient script. Only the name, hairstyle, genitals, and chest anatomy of the patients were different. Female-presenting faculty facilitators voiced the female patients and male-presenting faculty facilitators voiced the male patients. Of the 94 male students, 42 (44.7%) saw a male patient and 52 (55.3%) saw a female patient. Of the 93 female students, 45 (48.4%) saw a male patient and 48 (51.6%) saw a female patient. The time for each simulation was capped at 15 minutes regardless of resolution. The only other person in the room during the simulation was a standardized actor in the role of the nurse. Student performance was recorded on a formative assessment instrument that documented the order of actions taken, the timing of the actions, the utilization of point-of-care ultrasound (POCUS), and a surgery consultation, after which the case ended. The number of minutes from the beginning of the case until the use of the POCUS served as a surrogate variable for diagnostic reasoning. The number of minutes between the beginning of the case and the surgical consultation call served as the variable indicating successful intervention. The study was approved by the institutional review board.

The data collected during the ruptured-AAA simulation was cleaned and transformed prior to analysis in SPSS (version 27). Time stamps were translated into minutes passed between case start and POCUS use, call to surgery, and case end. Missing time stamps for POCUS use and the call to surgery indicated that these actions were not performed, and a value of 17 (max value + 1) was assigned. Because the Shapiro-Wilk test of normality was significant for both POCUS use (p < .000) and call to surgery, variables (p < .000) were transformed into rank outcomes. We conducted a two-way analysis of covariance (ANCOVA) to compare the effects of the independent variables of patient gender and student gender on POCUS use (diagnostic reasoning) and surgery call (successful intervention); two-way analysis of variance (ANOVA) of the effects of patient gender and student gender on either the dependent variable of POCUS use or surgery call; and logistic regression on the variable of correct diagnosis (ruptured AAA) by patient gender or student gender.

## Results

Several assumptions were tested prior to analysis. Levene’s test of equality was not significant for either ranked outcome i.e. call for surgical consultation (*F*(1, 185)= .118, p = .731) or POCUS use and (*F*(1, 185)= .126, p = .723) so the assumption of homogeneity of variance was satisfied. There was a linear relationship between the two dependent covariates, POCUS use and call to surgery, for each independent variable (patient gender and student gender). The Levene’s test of equality of variance was not significant for the factorial ANCOVA (p = .117), the two-way ANOVA of POCUS use (p = .908), or the two-way ANOVA of the call to surgery (p = .136).

In general, there were no significant interaction effects identified through the analyses. In the two-way ANCOVA of the time to surgery call with the time to POCUS use as the covariate, there was no significant interaction between the gender of the student and the gender of the patient (p = .819). There was also no significant interaction (p = .089) between student gender and patient gender in the factorial ANOVA of the surgery call. There was no significant effect of either the patient gender or the student gender alone on either the surgery call (p = .568 and p = .235, respectively) or POCUS use (p = .653 and p = .897, respectively). There was no significant difference in correct diagnosis identification between male and female students (p = .653).

Likewise, there was no significant difference in the correct diagnosis of either male or female patients (p = .210). However, there was one significant result: factorial ANOVA of POCUS use revealed a significant interaction (p = .010) between the gender of the student and the gender of the patient (Figure 1). *F*(1, 183) was 6.862 with an effect size of 0.36, indicating that 3.6% of the variance in time to POCUS was accounted for by the combined effect of patient and student gender.

**Figure 1.**
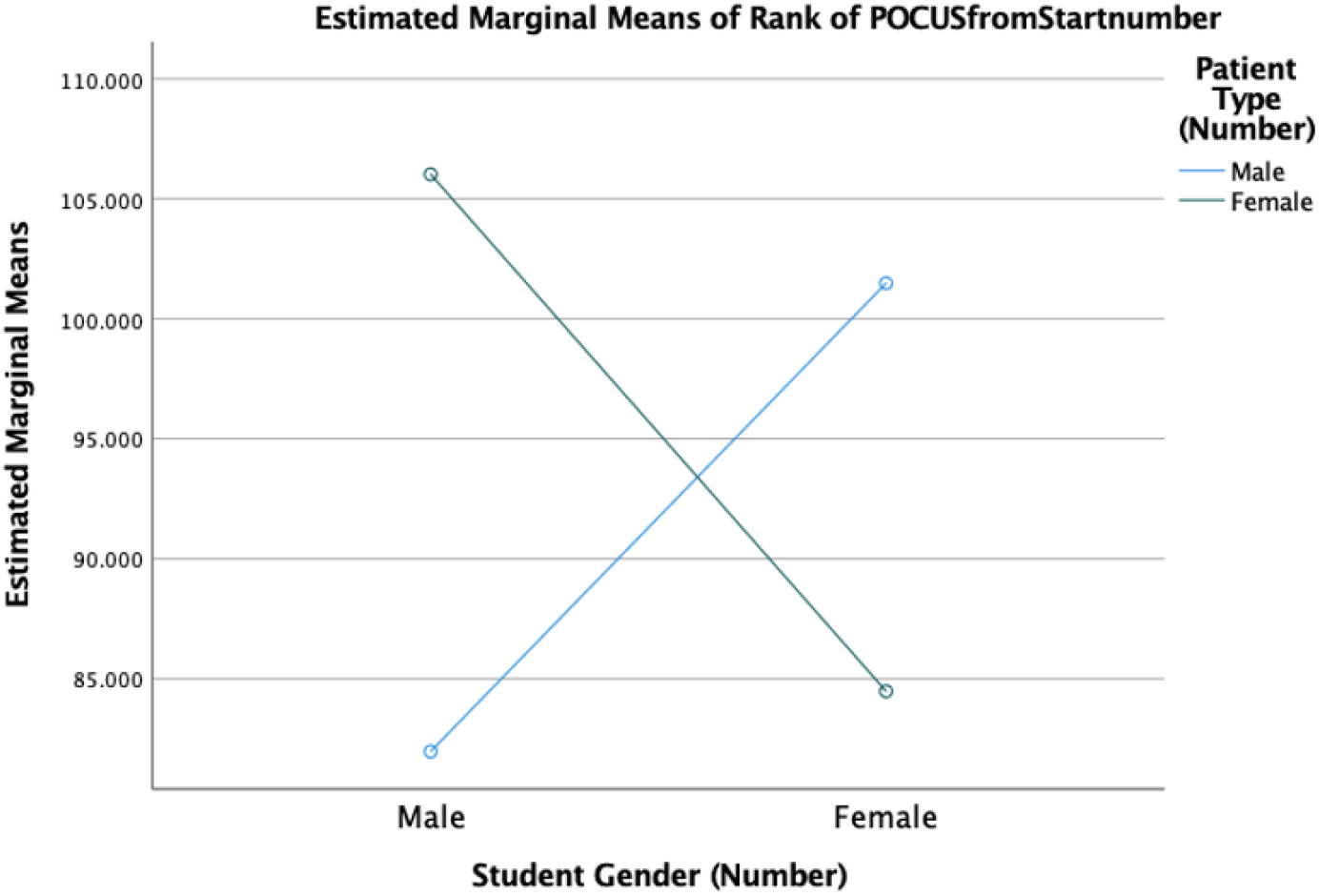

## Discussion

Simulation activities provide an opportunity to evaluate medical knowledge, clinical skills, and medical decision-making. Their reliability in replicating standardized clinical scenarios for multiple students makes them an effective tool to investigate learner reactions and judgments while controlling for specific variables. However, in a review of the literature, there is a paucity of studies that leverage clinical simulation to evaluate gender biases in medical education. This study sought to investigate the first possible setting in which gender bias in cardiac disease diagnosis and treatment may emerge for medical students; specifically, during the clerkship phase of their clinical education.

The use of POCUS served as a surrogate for diagnostic reasoning due to its ability to demonstrate the thought process of students, as corroborated by Kiesewatter et al. who categorized ultrasound as a strategic dimension of diagnostic knowledge^16^. Interestingly, while variance was observed in the time to initially reach for the POCUS between the two groups of students, there was no difference in the diagnostic accuracy among student or patient gender groups. From a treatment perspective, the time it took for students to call for the surgical consultation for the ruptured AAA was similar between both groups. In other words, once the students made the diagnosis, they reacted by consulting a surgeon, with no significant difference in the time it took to request this treatment; yet there was a statistical difference in the length of time it took to reach the diagnosis, which suggests slight in-group bias (3.6%). The noted difference in time to POCUS use between male and female students indicates a disparity in how students approach patients of a gender different from their own. The finding that female students were more quick to do ultrasounds on female patients suggests that diagnostic patient-physician concordance may begin to emerge sometime during practitioners’ medical education.

Patient-physician concordance is a key concept in healthcare delivery, as exemplified by a statistically significant difference in mortality associated with treatment of myocardial infarctions when the patient and provider were of the same gender^4^.

Despite the slight bias in imaging, students predominantly made accurate diagnoses and called for the correct intervention regardless of their own or their patient’s gender. This suggests that it is unlikely that medical students develop gender biases serious enough to impact clinical outcomes during the pre-clerkship/clerkship portion of their medical education, and that these heuristics may be formed later in training i.e. internship or residency. This possibility should be further investigated via a longitudinal study comparing outcome differences between new graduates and career physicians when treating male versus female patients^17^. Exploring these relationships would elucidate the exact stage in medical education and/or training where biases are most likely to develop. This knowledge could then be used to inform the development and integration of additional interventions to promote equitable care delivery at the appropriate curricular stage.

When addressing biases in care, the essential initial step is to acknowledge that bias, whether implicit or not, exists^18^. Individuals of all professions resort to pattern recognition to quickly and efficiently solve problems. However, this mindset poses dangers in the field of medicine due to its potential to perpetuate stereotypes and lead to compromised health outcomes. Studies show that repeated exposure and practice with various clinical and/or simulated scenarios are two important approaches to enlist in the battle against gender bias in the delivery of care. Actively using systemic reasoning skills and deliberate, slow thinking may help mitigate the effect of bias and ensure equitable care as well^18-20^.

### Limitations

The simulations were run using high-fidelity mannequins, which were equipped with features such as wigs, breasts, and female genitals to represent female patients. Additionally, a female facilitator provided the patient voice during the encounter. Studies have shown that high-fidelity mannequins are typically associated with improved student confidence and perception of simulation experiences^21^. As such, they have been considered an effective teaching tool to teach and assess a variety of professional and clinical skills including cardiovascular life support and trauma management, among others^22^. However, even with the aforementioned attempts to make the female simulated patients as realistic as possible, it is possible that the learners’ belief of the mannequin’s gender may have impacted how they interacted with and treated the patient. In addition, the simulations were timed, meaning that students were limited in the amount of time they could dedicate to diagnostic and/or treatment decision-making. Given unlimited time, students may have acted or performed differently. In addition, while the student groups were randomized, they were not equally distributed due to the limited availability of female facilitators to voice the mannequin. Lastly, since the study was performed at a single institution, replicating it at other institutions may allow for broader generalizability. Additional studies are needed to determine whether implicit bias against women affects other aspects of the diagnosis and treatment of cardiovascular disease.

## Conclusion

Data analysis revealed a slight, but statistically significant in-group bias in the time that it took to use POCUS, which was a surrogate for diagnostic reasoning. However, there was no significant difference between male and female learners in ultimately accurately diagnosing the AAA or calling for a surgery consultation. Likewise, the gender of the patient did not affect the accuracy of the diagnosis or treatment. It can thus be deduced that while gender bias affects imaging utilization decision-making early in medical education, prejudices leading to differential treatment outcomes for cardiovascular disease may emerge at a later time in training.

Given the repercussions of these biases, it is critical to investigate the stage during which they develop and institute ways to mitigate them. We believe that the success of simulation in fostering cultural competence and proficiency in procedural treatment can be extended to the setting of gender bias^23^. Implementing a more systematic approach to care in which students are trained to recognize nuance in patient presentation will reduce systemic bias’ detrimental effects on healthcare outcomes.

## Data Availability

The datasets generated and analyzed during the study are not publicly available but may be available from the corresponding author on reasonable request.

## Acknowledgments

The study was supported, in part, by the Dean of the Medical College of Georgia through the medical scholars program.

